# Effects of Weighted Seated Core Exercises on Immediate Postural Stability in Healthy Young Adults: An Exploratory Study

**DOI:** 10.64898/2026.11.03.26366886

**Authors:** Fumitaka Fujino, Masashi Yamakawa, Tatsuya Kono, Tsutomu Higaki, Kaoru Maeda

## Abstract

**Objectives:** This exploratory study investigated whether low-load, seated core exercises could produce immediate improvements in standing balance among healthy young adults, aiming to provide a foundation for safe, early-stage rehabilitation protocols.

**Methods:** Ten healthy males performed seated core exercises consisted of alternating hip flexion 5 times per side while seated and maintaining bilateral shoulder flexion at 120° shoulder flexion under four load conditions: upper-limb weight only, a 400-g empty box, a 1-kg ball, and a 2-kg ball. Immediately following the exercise, participants performed a 10-s single-leg stance. Transversus abdominis (TrA) thickness was measured via wireless ultrasound at baseline, post-exercise, and during the stance. Center of pressure (CoP) sway was recorded concurrently to evaluate postural stability.

**Results:** Left-side TrA thickness during stance increased significantly under the 1-kg and 2-kg load conditions compared to baseline (p < 0.05), although substantial missing data for the left-side measurements limited the robustness of this finding. No significant load-dependent effects were observed for CoP sway parameters (p > 0.05). A moderate positive correlation (ρ = 0.43, p = 0.002) was found between right-side TrA thickness and CoP sway velocity during the single-leg stance.

**Conclusions:** Low-load, seated core exercises do not yield immediate enhancements in standing stability in healthy populations. The results suggest that increased TrA thickness during a balance task may correlate with heightened postural instability rather than improved balance control. Further research is needed to determine whether these findings translate to clinical populations with impaired physical fitness.

## 1. Introduction

Promoting independence in activities of daily living (ADL) is a primary objective in rehabilitation for individuals with decreased physical fitness, such as older adults and patients in the early postoperative phase. Improvements in ADL are strongly influenced by standing balance^1)^. Strengthening the abdominal and lumbodorsal muscle groups (core muscles) is considered an effective intervention^2-4)^. Long-term core exercise programs that are typically performed multiple times per week are known to reduce fall risk^5)^ and increase physical activity^6)^; however, very few studies have investigated the immediate effects of such exercises^7-9)^. If standing balance could be improved immediately through low-risk exercises, it would facilitate the design of more effective staged rehabilitation programs for those with limited physical capacity.

The immediate effects of core exercises on standing balance remain controversial. Kaji et al.^9)^ reported a decrease in mediolateral center of pressure (CoP) sway during quiet standing with eyes closed after participants performed elbow-toe (a plank posture supported on elbows and toes) and hand-heel (a bridge-like posture supported on hands and heels) exercises for 30 s each. Conversely, some reports have found no immediate improvement in standing balance. Kadri et al.^8)^ had participants practice the abdominal drawing-in maneuver to induce contraction of the transversus abdominis (TrA) at a moderate intensity (based on the Borg CR-10 scale), after which they performed postural control tasks, such as single-leg stance or semi-tandem standing, while maintaining the abdominal drawing-in for 30 s. Their results showed no improvement of standing balance associated with the abdominal drawing-in maneuver. Furthermore, de Oliveira et al.^7)^ had participants perform 10 repetitions of 10-s holds for curl-ups, bridges, lateral planks, prone planks, and trunk anti-rotation exercises; however, no changes in CoP sway area or sway velocity were observed during single-leg stance and seated balance tasks. Thus, there is no consensus among researchers regarding whether core exercises immediately improve standing balance.

When interpreting the absence of immediate improvements in standing balance following core exercises, the studies of Kadri et al.^8)^ and de Oliveira et al.^7)^ shared two commonalities: (1) the participants were healthy young adults with no motor impairments, and (2) the exercise intensity was low or the duration/number of repetitions was small. However, if these exercises were applied to individuals with decreased physical fitness, the intensity and duration of the core exercises used by de Oliveira et al.^7)^ would presumably be considered excessive. Moreover, Kadri et al.^8)^ attempted to maintain the core muscles in a contracted state by having participants perform an abdominal drawing-in maneuver. Because this maneuver requires practice to maintain consistently, it would likely be difficult to apply to individuals with a diminished capacity for learning new motor skills. Risk management during exercise is crucial in rehabilitation for individuals with decreased physical fitness. In an acute setting, such as the early postoperative phase, it is recommended to progress exercises in stages, starting from movements that can be performed in bed and advancing gradually to light walking^10,11)^. During this phase, it is vital that patients become capable of transferring and walking as early as possible without risking a fall. Therefore, it is important to investigate the effects of exercises configured with appropriate intensities, durations, and postures that can be safely performed in individuals with decreased physical fitness (e.g., in a seated position) while aiming to immediately improve standing balance. Such investigations may provide a foundation for rehabilitation interventions that balance safety and efficacy.

To investigate the extent to which core muscle activity contributes to improvements in standing balance, it is critical to determine whether core exercises facilitate core muscle activation using the most direct indices possible. In previous studies evaluating the immediate effects of core exercises^7-9)^, muscle activity during the subsequent standing balance task was not measured; thus, the relationship between core muscle activation and standing balance remains unclear. Direct measures of core muscle activity include surface electromyography (EMG)^8,12)^ and ultrasonography (US)^4)^.

While EMG is widely used, its ability to isolate deep muscles such as the TrA is limited by signal crosstalk from superficial muscles. Ultrasound imaging offers high spatial resolution suitable for isolating these deep contractions. Furthermore, utilizing a wireless ultrasound probe fixed to the trunk minimizes mechanical interference during single-leg stance tasks compared to traditional wired devices.

The purpose of this study was to explore whether low-load, seated core exercises immediately improve standing balance in healthy young adults, focusing on the TrA, which plays a vital role in stabilizing the pelvis and lumbar spine^4,13)^. Specifically, we established three objectives: to determine the degree to which low-load, seated core exercises facilitate TrA activation, to identify the correlation between changes in muscle thickness and CoP sway, and to clarify how different load conditions influence these parameters. To our knowledge, this is the first study to evaluate TrA thickness during a standing balance task while investigating the immediate effects of a low-load exercise intervention.

## 2. Methods

### 2.1. Participants

The participants were 10 males (mean age: 20.1 years, SD=0.3, CV=1.6; mean height: 171.5 cm, SD=4.6, CV=2.7; mean weight: 65.3 kg, SD=5.6, CV=8.6). None of the participants had a history of orthopedic or neurological disorders. The participants received written explanations regarding the study and provided written consent. This study was approved by the Research Ethics Committee of Morinomiya University of Medical Sciences (Approval Number: 2024-032). Each participant’s non-dominant leg was designated as the supporting leg for maintaining the single-leg stance. The non-dominant leg (1 right, 9 left) was defined via interview as the leg contralateral to the one preferred for kicking a ball.

### 2.2. Apparatus

CoP sway during the single-leg stance was recorded using a force plate (East Medic Co., Ishikawa, Japan) comprising four pressure sensors (LCB-1KN; NTS Co., Ltd., Tokyo, Japan). Signals from the pressure sensors were fed into an arithmetic circuit (Electro Design Co., Chiba, Japan) that calculated the CoP position signals along the anteroposterior and mediolateral axes. The signals were input to a PC via an A/D converter (AIO-163202FX-USB, Contec Co., Osaka, Japan) at a sampling frequency of 2000 Hz and a 16-bit quantization resolution. The data were stored on the hard disk of the PC as time-series data, using biological signal recording software (Vital Recorder; Kissei Comtec Co., Nagano, Japan). A visual target was placed 1.5 m directly in front of the leading edge of the force plate at a height of 1.5 m to serve as a fixation point during single-leg standing. To cue the movement timing during the core exercises, an iOS application (Smart Metronome & Tuner, Version 4.1.2, Tomohiro Ohata) was utilized to generate a rhythmic sound set to 30 beats/min (0.5 Hz).

To capture transverse cross-sectional images of the transversus abdominis immediately prior to the core exercises and during the single-leg stance, a wireless ultrasound imaging device (Vscan Air CL, GE Healthcare Co., Tokyo, Japan) was affixed to both sides of the participant’s abdomen using a custom-made fixation device constructed from Styrofoam and Velcro tape (Fig. 1) (Photographs depicting the experimental setup and specific exercise postures are available from the corresponding author upon reasonable request). The center of the linear probe was aligned with the intersection of the participant’s anterior axillary line and a horizontal line passing through the umbilicus. Probe positions were marked on the skin to monitor and correct any displacement during tasks. The imaging mode was B-mode, and the depth was set to either 6 cm or 7 cm depending on the participant.

**Figure 1.** Experimental tasks and ultrasound measurement of the transversus abdominis thickness.^*^ A: Seated core exercise task under the empty box load condition. B: Single-leg standing task with bilateral wireless ultrasound probes secured to the abdomen using a custom fixation device. C: Representative transverse ultrasound image of the left transversus abdominis (TrA). The distance between the tips of the two white arrows indicates TrA thickness. ^*^Photographs depicting the experimental setup and specific exercise postures are available from the corresponding author upon reasonable request.

### 2.3. Procedure

#### 2.3.1. Core Exercise

The core exercise consisted of alternating hip flexion to lift the soles of the feet off the floor for 10 s (5 times per side) while seated and maintaining bilateral shoulder flexion at 120° (Fig. 1A) (Photographs depicting the experimental setup and specific exercise postures are available from the corresponding author upon reasonable request). To alter the load conditions applied to the core muscles, participants performed the exercise under four conditions, while sequentially holding the following three objects with both hands: an empty box (15 cm × 25 cm × 10 cm, 400 g), a 1-kg medicine ball, and a 2-kg medicine ball (approx. 25 cm in diameter; Systemstyle Co., Gunma, Japan). An exercise trial was also conducted without holding any object.

#### 2.3.2. Single-leg Standing

Participants crossed their arms in front of the chest and maintained a single-leg stance on the non-dominant leg for 10 s, keeping the dominant foot off the floor by flexing the knee (Fig. 1B) (Photographs depicting the experimental setup and specific exercise postures are available from the corresponding author upon reasonable request). During the task, participants were instructed to gaze at the anterior fixation point.

#### 2.3.3. Measurement Order

The order of measurement was as follows. First, the participants held a single-leg stance for 10 s. Approximately 5 s after initiation of the single-leg stance, one ultrasound image was captured for each side. Imaging was executed by two examiners, positioned on each of the left and right side, who concurrently tapped the capture button on the smartphone application used to control the probes. The timing for capturing the images was coordinated as follows: the left-side examiner started a stopwatch exactly when the participant initiated the single-leg stance, and when the stopwatch displayed 5 s, the left-side examiner signaled the right-side examiner. Center of pressure sway and TrA muscle thickness measured during this initial stance were defined as baseline values during single-leg stance. Subsequently, one trial of single-leg stance was performed immediately after the core exercise under each load condition, and an ultrasound image was captured for each side approximately 5 s after initiation of the stance. The load conditions were applied in the following order: upper limb elevation only, holding an empty box, holding a 1-kg medicine ball, and holding a 2-kg medicine ball. The object weights were sequenced in ascending order to prevent fatigue from heavy loads from influencing the effects observed under lighter load conditions.

#### 2.3.4. Attachment of the Fixation Device and Ultrasound Probe, and Assessment of Intra-rater Reliability

Across all participants, the same examiner was responsible for the fixation and adjustment of the probe on one side of the participant’s trunk (TH for the right side, KM for the left side). The following procedure was performed to evaluate intra-rater reliability of probe fixation. One US image was taken immediately after securing the probe to the seated participant’s body. The outline of the fixation device in contact with the skin was then marked on the skin using a pen, and the fixation device and probe were removed as a single unit. The probe was reattached with the fixation device aligned with the marker lines, and a second US image was captured (Fig. 1C) (Photographs depicting the experimental setup and specific exercise postures are available from the corresponding author upon reasonable request). Intra-rater reliability was assessed using the TrA muscle thickness values measured from these two images. The muscle thickness value obtained from the second image was utilized as the baseline reference value prior to the core exercises (i.e., just before each core exercise with each load).

#### 2.3.5. Visual Confirmation and Adjustment of Fixation Device Position During Ultrasound Imaging

During the measurements for every participant, the examiners visually verified the positional relationship between the fixation device and the markers prior to both the core exercise and the single-leg stance. If an examiner determined that the fixation device had shifted from the marker line, the examiner manually corrected its position.

### 2.4. CoP Signal Processing and Muscle Thickness Measurement

CoP signals were resampled to 100 Hz using BIMUTAS II software (Kissei Comtec Co., Nagano, Japan). The mean velocity of CoP sway (cm/s) and standard deviation (SD) of CoP displacement in the anteroposterior and mediolateral directions (cm) were calculated using custom Excel VBA code in Microsoft Excel (Microsoft Corp., Redmond, WA, USA). TrA thickness was measured from the US images as the distance between the borders of the muscle fibers at the image midline (Fig. 1C) (Photographs depicting the experimental setup and specific exercise postures are available from the corresponding author upon reasonable request).

### 2.5. Statistical Analysis

Statistical analysis was performed using jamovi (version 2.3). Normality was assessed with the Shapiro–Wilk test. Given the exploratory nature of the study and the potential for non-normal distribution in a small sample (n=10), the effects of load conditions were evaluated using the Friedman test. When significant effects were detected, post-hoc comparisons were conducted using the Durbin–Conover method. The relationship between muscle thickness and CoP parameters was analyzed using Spearman’s rank correlation coefficient. For all tests, the significance level was set at p < 0.05.

## 3. Results

Of the ultrasound images captured, TrA thickness could not be measured in 7 out of 100 images due to poor image quality. Image quality issues were more frequent on the left side (where 5 images were unmeasurable before the exercise and 2 during the stance) whereas no images on the right side were excluded. Consequently, data from 7 participants were included in the longitudinal analysis of muscle thickness.

Intra-rater reliability of the probe fixation differed by side: a significant and strong correlation between initial and refixation measurements was found only on the right side (ρ=0.94, p<0.001). On the left side, the correlation was lower and non-significant (ρ=0.57, p=0.11). Regarding TrA thickness before the exercise, no significant effect of load condition was observed (Table 1). However, during the single-leg stance, a significant effect of load condition was observed only on the left side (χ2=11.6, df=4, p=0.02) (Table 1). Post-hoc tests indicated that muscle thickness was significantly greater under the 1-kg and 2-kg conditions compared to baseline (p<0.05), and significantly greater under the empty box, 1-kg, and 2-kg conditions compared to body weight alone (p<0.05) (Table 1). A direct comparison between muscle thickness before the exercise and during the single-leg stance was not performed due to the fundamental differences in posture (seated vs. standing).

**Table 1.** Effects of core exercise on muscle thickness and CoP parameters across different load conditions.

| Category | Parameter | Side | Baseline | UL elevation | 400-g box | 1-kg ball | 2-kg ball | Friedman test (p) |
|---|---|---|---|---|---|---|---|---|
| Muscle thickness (mm)<br>(n=7) | Before exercise | Right | 39 (8) | 39 (3) | 38 (7) | 38 (9) | 41 (12) | 0.92 |
|  |  | Left | 37 (15) | 34 (7) | 34 (6) | 35 (7) | 35 (10) | 0.93 |
|  | During SLS | Right | 41 (3) | 39 (7) | 41 (6) | 40 (8) | 41 (9) | 0.97 |
|  |  | Left | 38 (9) | 38 (9) | 40 (12)** | 42 (9)*,** | 42 (11)*,** | 0.02 |
| CoP<br>(n=10) | Mean velocity (cm/s) | - | 5.1 (1.1) | 4.6 (1.2) | 4.7 (1.3) | 4.6 (1.3) | 4.4 (1.2) | 0.08 |
|  | SD a–p (cm) | - | 7.1 (2.5) | 8.1 (3.4) | 9.4 (3.2) | 7.7 (2.2) | 7.8 (2.4) | 0.95 |
|  | SD m–l (cm) | - | 6.0 (1.7) | 6.5 (1.7) | 6.2 (2.7) | 6.3 (2.3) | 6.4 (1.8) | 0.39 |

Abbreviations: BL: baseline; CoP: center of pressure; SD a–p: standard deviation of CoP sway in the anteroposterior direction; SD m–l: standard deviation of CoP sway in the mediolateral direction; SLS: single-leg stance; UL: upper-limb
\* significantly different from baseline (p<0.05, Durbin–Conover test); \*\* significantly different from UL elevation (upper-limb weight alone) (p<0.05, Durbin–Conover test).

Standing balance parameters showed no significant influence from the core exercise load conditions (Table 1). There were no significant changes in the mean velocity of CoP sway (χ2=8.24, df=4, p=0.08), mediolateral SD (χ2=0.72, df=4, p=0.95), or anteroposterior SD (χ2=4.16, df=4, p=0.39). A moderate positive correlation was identified between right-side muscle thickness and the mean velocity of CoP sway during the single-leg stance (ρ=0.43, p=0.002) (Table 2).

**Table 2.**
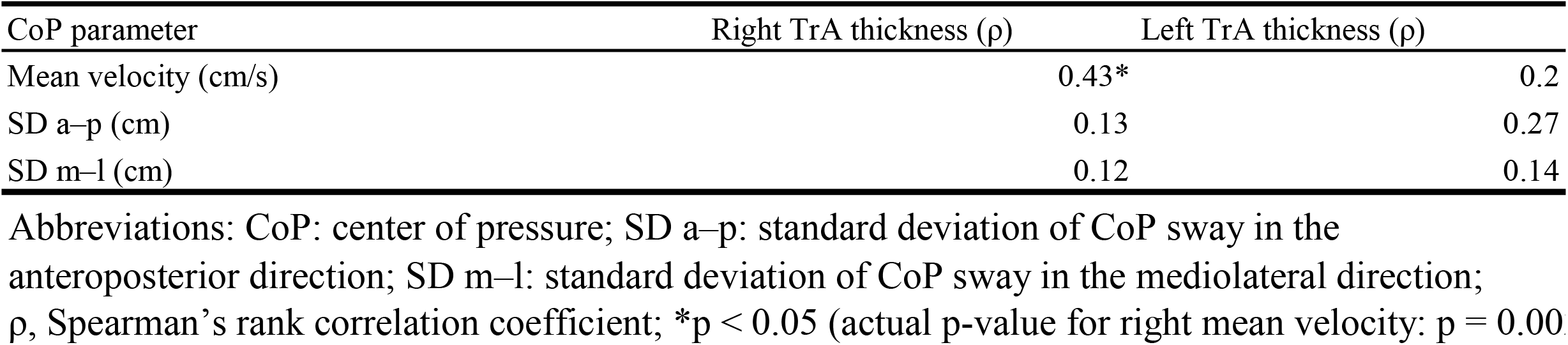
Spearman’s rank correlation coefficients (ρ) between TrA thickness and CoP parameters during single-leg standing (n = 10)

| CoP parameter | Right TrA thickness (ρ) | Left TrA thickness (ρ) |
|---|---|---|
| Mean velocity (cm/s) | 0.43* | 0.2 |
| SD a–p (cm) | 0.13 | 0.27 |
| SD m–l (cm) | 0.12 | 0.14 |

Abbreviations: CoP: center of pressure; SD a–p: standard deviation of CoP sway in the anteroposterior direction; SD m–l: standard deviation of CoP sway in the mediolateral direction; ρ, Spearman’s rank correlation coefficient; \*p < 0.05 (actual p-value for right mean velocity: p = 0.00[illegible]

## 4. Discussion

This exploratory study investigated whether low-load, seated core exercises immediately improve standing balance while simultaneously assessing TrA thickness during the task. No immediate improvement in standing stability was observed following the intervention. Instead, a positive correlation between right-side TrA thickness and CoP sway velocity suggested that greater TrA thickness during the balance task was associated with poorer stability.

The participants were healthy young men with relatively small variations in physical characteristics (CV=1.6-8.6). While this provided a stable baseline, the study did not assess individual physical fitness parameters, such as muscle strength or baseline balance capacity, which may have influenced the outcomes. The small sample size (n=10) represents a major limitation of this exploratory study, and future research should include a larger, more diverse population to improve generalizability.

A significant increase in TrA thickness was observed primarily on the left side, suggesting that the hip-flexion task might not have adequately facilitated bilateral core muscle activation. Kadri et al.^8)^ found no immediate improvement using abdominal drawing-in maneuvers and speculated that higher-intensity contractions might be necessary. However, if the load is excessively high, increased activation of superficial global muscles could disrupt the segmental stabilization provided by deep local muscles such as the TrA^14)^. Future investigations should determine optimal exercise loads based on individual physical fitness characteristics rather than fixed weights.

We used CoP sway during a single-leg stance, which is widely utilized for static balance evaluation, as the primary outcome measure^4,7,8)^. However, repeatedly performing the task exclusively on the non-dominant leg may have induced localized muscle activation. As 90% of participants used their left leg for support, this repetition likely contributed to the significant muscle thickness increase observed only on the left side. Furthermore, static balance assessment may not capture the dynamic balance demands associated with core muscle function. Future studies should incorporate outcome measures such as balance response speed and performance on unstable surfaces to better replicate ADL demands.

While high intra-rater reliability was achieved for probe fixation on the right side, lower reliability on the left side necessitates caution in interpreting the bilateral data. Refining the fixation method, perhaps through 3D-printed custom devices or additional anatomical markers, is crucial for future research. In addition, measuring thickness at a single point may not reflect the functional behavior of the entire TrA. Future measurements should utilize techniques with improved temporal and spatial resolution to capture the complex dynamics of deep trunk muscle activity during movement.

A moderate positive correlation (ρ=0.43) between right-side muscle thickness and mean CoP sway velocity was a key finding. This contrasts with the general hypothesis that core contraction increases trunk stiffness to enhance stability, implying instead that increased thickness might be associated with impaired postural control. Several mechanisms may explain this: inappropriate contraction directions inducing joint perturbation^15)^, excessive contraction hindering the co-activation of antagonist muscles^16)^, or the localized activation of one muscle disrupting the integrated function of the entire core^17)^. While we sequenced loads to minimize fatigue, we cannot rule out its influence on balance. Sufficient rest periods and varied exercise methodologies are needed to isolate the effects of load from muscle fatigue.

In conclusion, this study demonstrated that low-load, seated core exercises do not yield an immediate improvement in standing stability among healthy young adults. Furthermore, the observed correlation suggests that an increase in TrA muscle thickness may not necessarily contribute to enhanced stability, and may instead disrupt postural control. However, since this was an exploratory pilot study with a small sample size (n=10), these findings should be interpreted with caution. Further research with a larger, more diverse population is needed to clarify the relationship between core muscle activity and functional balance.

## Data Availability

All data produced in the present study are available upon reasonable request to the authors

